# Sinonasal disease among patients with primary ciliary dyskinesia – an international study

**DOI:** 10.1101/2022.11.23.22282583

**Authors:** Yin Ting Lam, Jean-François Papon, Mihaela Alexandru, Andreas Anagiotos, Miguel Armengot, Mieke Boon, Andrea Burgess, Suzanne Crowley, Sinan Ahmed. D. Dheyauldeen, Nagehan Emiralioglu, Ela Erdem Eralp, Christine van Gogh, Yasemin Gokdemir, Onder Gunaydın, Eric G. Haarman, Amanda Harris, Isolde Hayn, Hasnaa Ismail-Koch, Bülent Karadag, Céline Kempeneers, Sookyung Kim, Philipp Latzin, Natalie Lorent, Ugur Ozcelik, Charlotte Pioch, Anne-Lise ML Poirrier, Ana Reula, Jobst Roehmel, Panayiotis Yiallouros, the EPIC-PCD team, Myrofora Goutaki

**Affiliations:** Institute of Social and Preventive Medicine, University of Bern, Switzerland; Graduate School for Cellular and Biomedical Sciences, University of Bern, Bern, Switzerland; Assistance Publique-Hôpitaux de Paris (AP-HP), Université Paris-Saclay, Hôpital Bicêtre, Service d’ORL, Le Kremlin-Bicêtre, France; Faculté de Médecine, Université Paris-Saclay, Le Kremlin-Bicêtre, France; Department of Otorhinolaryngology, Nicosia General Hospital, Nicosia, Cyprus; Department of Otorhinolaryngology, and Primary Ciliary Dyskinesia Unit, La Fe University and Polytechnic Hospital, Valencia, Spain; Medical School, Valencia University, Valencia, Spain; Department of Paediatrics, University Hospital, Leuven, Belgium; Primary Ciliary Dyskinesia Centre, Southampton Children’s Hospital, Southampton NHS Foundation Trust, Southampton, UK; Paediatric Department of Allergy and Lung Diseases, Oslo University Hospital, Oslo, Norway; Department of Otorhinolaryngology, Head and Neck Surgery, Oslo University Hospital, Oslo, Norway; Faculty of Medicine, University of Oslo, Norway; Department of Pediatric Pulmonology, Hacettepe University, School of Medicine, Ankara, Turkey; Department of Pediatric Pulmonology, Marmara University, School of Medicine, Istanbul, Turkey; Department of Otorhinolaryngology - Head and Neck Surgery, Amsterdam UMC, Amsterdam, The Netherlands; Department of Otorhinolaryngology, Hacettepe University, School of Medicine, Ankara, Turkey; Department of pediatric pulmonology, Emma Children’s Hospital, Amsterdam UMC, Vrije Universiteit, Amsterdam, The Netherlands; Southampton Children’s Hospital, University of Southampton, Southampton, UK; Primary Ciliary Dyskinesia Centre, NIHR Respiratory Biomedical Research Centre, University of Southampton, Southampton, UK; Department of Otorhinolaryngology, Head and Neck Surgery, Charité-Universitätsmedizin Berlin, Berlin, Germany; Division of Respirology, Department of Pediatrics, University Hospital Liège, Liège, Belgium; Division of Paediatric Respiratory Medicine and Allergology, Department of Paediatrics, Inselspital, University Hospital, University of Bern, Switzerland; Department of Respiratory Diseases, University Hospital, Leuven, Belgium; Department of Pediatric Pulmonology, Immunology and Critical Care Medicine, Charité-Universitätsmedizin Berlin, Berlin, Germany; Department of Otorhinolaryngology, University Hospital Liège, Liège, Belgium; Biomedical Sciences Department, CEU-Cardenal Herrera University, Castellón, Spain; Molecular, Cellular and Genomic Biomedicine Group, IIS La Fe, Valencia, Spain; Medical School, University of Cyprus, Nicosia, Cyprus; Pediatric Pulmonology Unit, Hospital ‘Archbishop Makarios III’, Nicosia, Cyprus

**Keywords:** Primary ciliary dyskinesia, epidemiology, orphan pulmonary diseases, sinusitis, upper airway, QoL (upper airway system)

## Abstract

**Background:** Although most patients with primary ciliary dyskinesia (PCD) report sinonasal symptoms, little is known about symptom frequency and severity. We describe sinonasal manifestations among PCD patients using data from the Ear, nose, and throat (ENT) Prospective International Cohort of PCD patients.

**Methods:** We included data from participants with routine clinical ENT examinations and complete FOLLOW-PCD symptoms questionnaires from the same visit or within 2 weeks. We compared the prevalence, reported symptoms, and clinical findings among children and adults and identified potential factors associated with increased risk of sinonasal disease using ordinal regression.

**Results:** We included 397 (53% male) participants from 12 centres with median age 15 years (IQR 9– 22). Almost all (352; 89%) reported chronic nasal symptoms. More adults (34; 26%) than children (11; 5%) reported anosmia or hyposmia. Among 140 participants who completed SNOT-22 questionnaires, quality of life was moderately affected by sinonasal symptoms (median score 31; IQR 22–45). We observed nasal polyps among 52 (15%) of 352 participants and hypertrophic inferior nasal turbinates among 129 (34%) of 353 participants; facial pain was recorded among 51 (13%) of 353 participants. More adults than children had nasal polyps, hypertrophic turbinates, deviated septum, and facial pain. We found age 10 and older the only factor associated with increased risk of sinonasal disease.

**Conclusions:** Our study reinforces the importance of regular sinonasal evaluations for patients of all ages with PCD and the need for developing evidence-based guidelines for sinonasal treatments as part of overall PCD management.

## Introduction

Sinonasal symptoms among patients with primary ciliary dyskinesia (PCD) are as common as lower respiratory symptoms [1, 2]. Often present from birth, rhinitis is one of the first signs of PCD and usually persists throughout life [3–6]. With impaired respiratory ciliary movement and reduced mucociliary clearance, nasal secretions depend only on gravity and airflow transport [1, 7]. Sinonasal problems may manifest with rhinorrhea or blocked nose, facial pain, and headaches [8, 9]. With PCD, symptoms are part of daily life, often considered normal, and likely underreported during routine consultations. Sinonasal disease is also characterised by recurrent upper respiratory infections, often leading to chronic rhinosinusitis (CRS). Despite the clinical burden, sinonasal manifestations are frequently neglected and in many centres, ear-nose-throat (ENT) assessments are not part of routine multidisciplinary PCD care, particularly for adults [10–16]. Since sinuses may function as bacterial reservoirs for pulmonary infections later leading to lung function impairment, sinus infections are often considered only after unsuccessful treatment of pulmonary infections [17–22]. Encountered among 15–30% patients with PCD compared with 3–4% among the general population, nasal polyps are also common [6, 24]. Other sinonasal manifestations among patients with PCD include hypoplasia or agenesis of paranasal sinuses [8, 23].

The few published studies on sinonasal manifestations in PCD are mostly retrospective, include small numbers of 20–60 participants who are primarily children, and obtain data from chart reviews where symptoms were collected in a nonstandard way [25, 2]. Little is known about progression of sinonasal disease with age or with increased frequency of sinonasal symptoms. We aimed to describe the prevalence of patient-reported sinonasal symptoms and sinonasal examination findings among children and adults with PCD and identify possible risk factors associated with sinonasal disease.

## Methods

### Study design and population

Our study analyses cross-sectional baseline data from the ENT prospective, international cohort of patients with PCD (EPIC-PCD)—the first PCD cohort focused on upper airway disease manifestations [26]. We set up EPIC-PCD in February 2020 to follow PCD patients at their routine ENT consultations. Participants did not undergo additional testing for our study purposes. EPIC-PCD is hosted at the University of Bern (clinicaltrials.gov identifier: NCT04611516). For our collaborative study, 12 participating centres (Amsterdam, Ankara, Berlin, Bern, Cyprus, Istanbul, Leuven, Liège, Oslo, Paris, Southampton, Valencia) in 10 countries contributed data. For our analysis, we included data entered in the database by 31 July 2022 participants with PCD of all ages who underwent ENT examinations and completed symptoms questionnaires at the same visit or within 2 weeks.

We received ethical approval from all participating centers and human research ethics committees in accordance with local legislation. We obtained informed consent or assent from either participants or parents or caregivers of participants 14 years or younger. Our report conforms with the Strengthening the Reporting of Observational studies in Epidemiology (STROBE) statement [27].

### Patient-reported symptoms and quality of life

For collecting patient-reported symptoms, we used the disease-specific FOLLOW-PCD questionnaire (version 1.0), which is part of the FOLLOW-PCD form developed to collect clinical information for research and clinical follow-up in a standardised way [28]. There are age-specific versions of the FOLLOW-PCD questionnaire for adults, adolescents 14–17 years, and parents or caregivers of children with PCD 14 years and younger. The FOLLOW-PCD questionnaire is available in languages of participating centres. Sinonasal symptoms questions ask about frequency and characteristics of symptoms during the past three months, specifically focusing on chronic nasal symptoms, snoring, and headaches, as well as more frequent ENT symptoms during the past 12 months. Symptom frequency options included daily, often, sometimes, rarely, and never (five-point Likert scale). Lifestyle questions asked about smoking exposure and living conditions during the past 12 months. Depending on available response categories, we recoded missing answers as “unknown,” “no,” or “never.”

Based on local protocols, if distributed during the clinic visit we also collected information about quality of life (QoL) using the Sino-Nasal Outcome Test (SNOT-22) [29]. SNOT-22 is a validated CRS health-related QoL outcome measure. Participants score CRS-related items 0–5 each from “no problem” to “problem as bad as it can be”. In total, SNOT-22 ranges between 0–110; corresponding to a mild (0–20), moderate (21–50), or severe (≥ 51) effect of CRS on QoL.

### Sinonasal examinations

The EPIC-PCD is nested in routine care and follows participants at their usual ENT consultations. Performed by an ENT specialist according to local protocols, routine ENT consultations included clinical sinonasal examinations by nasal endoscopy or anterior rhinoscopy if tolerated by the participant. Examination findings were recorded in a standardised way using the ENT examination module of the FOLLOW-PCD form [28]. We recorded the proportion of the total nasal cavity volume occupied by nasal polyps using a semi-quantitative measure— the Lildholdt score— described as “partially blocking” (Lildholdt scores 1–2) and “fully blocking” (Lildholdt score 3) [30]. We recorded, reported, and present missing information from sinonasal examinations as missing.

### Diagnosis and other clinical information from charts

Participants were diagnosed according to European Respiratory Society (ERS) guidelines [31]. Definite PCD diagnosis was confirmed by presence of hallmark ultrastructural defects seen in transmission electron microscopy (TEM) or by identification of bi-allelic pathogenic mutations in PCD genes. Remaining participants were diagnosed by a combination of several tests, including nasal nitric oxide, high-speed video microscopy analysis, or immunofluorescence. We collected data on laterality defects from medical records, and when it was available, past medical history information particularly about neonatal rhinitis. Lastly, in addition to the basic dataset, some participating centres contributed information on prescribed sinonasal management. We entered all collected data in the study database—which uses the Research Electronic Data Capture (REDCap) software—based on the FOLLOW-PCD form [28].

### Statistical analysis

We described characteristics of the population, patient/parent-reported sinonasal symptoms, and sinonasal examination findings, for the total population and separately among age groups 0–6, 7–14, 15–30, 31–50, and 50 years and older. For continuous variables, we used median and interquartile range (IQR); for categorical variables, we used numbers and proportions, calculating Wilson 95% confidence intervals (CI) for proportions. We compared differences between age groups using Pearson’s Chi square, Wilcoxon rank-sum, and Kruskal-Wallis rank test. We created a composite outcome variable for sinonasal disease consisting of three variables: patient-reported headache while bending down as a proxy for sinusitis, ENT examination findings of nasal polyps, and facial pain. Each of them scored either 0 (absence) or 1 (presence). Total scores ranged from 0 to 3. We assessed factors possibly associated with sinonasal disease such as age, age of diagnosis, sex, study centre, smoking status of either active or passive smoke exposure, and season when ENT consultations occurred in a multivariable ordinal logistic regression model. We chose factors based on clinical importance and data availability. There was collinearity of age and age of diagnosis so it was not possible to include both in our main model; separate models showed similar results so we included age. After exploring linear and non-linear effects of age as continuous variable, we chose to include age by decades in the final model. We excluded study centre from the full model due to restricted sample size; however, we conducted sensitivity analyses with study centre alone and with age. Lastly, among a subgroup of participants with available TEM results, we repeated the model including age and category of ciliary ultrastructural defect to study if ciliary ultrastructural defect was associated with risk for sinonasal disease. We performed all analyses with Stata version 15 (StataCorp LLC, Texas, USA).

## Results

### Study population

By the end of July 2022, 448 (89%) of 505 invited patients with PCD enrolled in the EPIC-PCD cohort (figure 1). Of them, 397 (53% male) participants with median age 15 years (IQR 9–22) entered in the database fulfilled eligibility criteria for ENT consultation and completed FOLLOW-PCD questionnaire at the same visit or within 2 weeks (table 1). Among participants, 257 (65%) were children, 140 (35%) adults, and 142 (36%) had situs inversus totalis. With regards to participant diagnostic status, 252 (63%) had definite PCD diagnoses based on ERS guidelines [31] with a bi-allelic PCD-causing mutation or a hallmark defect identified by TEM (table S1); 125 (32%) participants were diagnosed by a combination of several other tests; and 21 (5%) were newly diagnosed participants with diagnostic results pending at enrolment.

**Table 1:** Characteristics of EPIC-PCD participants, overall and by age group (N=397)

|  | **Total**<br>**N (%)** | **Age 0-6 y**<br>**N (%)** | **Age 7-14 y**<br>**N (%)** | **Age 15-30 y**<br>**N (%)** | **Age 31-50 y**<br>**N (%)** | **Age >50 y**<br>**N (%)** | **p-value<sup>a</sup>** |
| --- | --- | --- | --- | --- | --- | --- | --- |
| **Number of participants** | 397 (100) | 44 (100) | 130 (100) | 157 (100) | 43 (100) | 23 (100) |  |
| Age, median (IQR) | 15 (9–22) | 4 (2–5) | 10 (8–12) | 16 (15–21) | 38 (34–43) | 57 (56–62) |  |
| Male sex | 211 (53) | 23 (52) | 72 (55) | 81 (52) | 24 (56) | 11 (48) | 0.937 |
| Age of PCD diagnosis,<br>median (IQR) | 9 (4–17) | 1 (0–2) | 6 (1–8) | 12 (8–17) | 34 (29–36) | 51 (43–55) |  |
| **Consanguinity** |  |  |  |  |  |  | 0.001 |
| Yes | 119 (30) | 7 (16) | 45 (35) | 52 (33) | 11 (26) | 4 (17) |  |
| No | 136 (35) | 13 (29) | 40 (30) | 61 (39) | 19 (44) | 3 (13) |  |
| Not reported | 142 (35) | 24 (55) | 45 (35) | 44 (28) | 13 (30) | 16 (70) |  |
| **Situs** |  |  |  |  |  |  | <0.001 |
| Situs inversus totalis | 142 (36) | 25 (57) | 46 (35) | 60 (38) | 7 (16) | 4 (17) |  |
| Situs ambiguous | 4 (1) | 0 (0) | 1 (1) | 3 (2) | 0 (0) | 0 (0) |  |
| Situs solitus | 245 (62) | 18 (41) | 82 (63) | 94 (60) | 32 (74) | 19 (83) |  |
| Not reported | 6 (1) | 1 (2) | 1 (1) | 0 (0) | 4 (9) | 0 (0) |  |
| **Cardiovascular malformation present** |  |  |  |  |  |  | 0.088 |
| Yes | 34 (9) | 7 (16) | 11 (8) | 14 (9) | 2 (5) | 0 (0) |  |
| No | 295 (74) | 30 (68) | 104 (80) | 114 (73) | 32 (74) | 15 (65) |  |
| Not reported | 68 (17) | 7 (16) | 15 (12) | 29 (18) | 9 (21) | 8 (35) |  |
| **Neonatal rhinitis** |  |  |  |  |  |  | 0.189 |
| Yes | 125 (32) | 15 (34) | 47 (36) | 49 (31) | 12 (28) | 2 (9) |  |
| No | 88 (22) | 7 (16) | 27 (21) | 39 (25) | 11 (26) | 4 (17) |  |
| Not reported | 184 (46) | 22 (50) | 56 (43) | 69 (44) | 20 (46) | 17 (74) |  |
| **Neonatal cough** |  |  |  |  |  |  | <0.001 |
| Yes | 129 (33) | 15 (34) | 44 (34) | 59 (38) | 10 (23) | 1 (4) |  |
| No | 91 (23) | 3 (7) | 31 (24) | 40 (25) | 14 (33) | 3 (13) |  |
| Not reported | 177 (44) | 26 (59) | 55 (42) | 58 (37) | 19 (44) | 19 (83) |  |
| **Neonatal respiratory distress** |  |  |  |  |  |  | 0.554 |
| Yes | 181 (46) | 26 (59) | 60 (46) | 67 (43) | 20 (46) | 8 (35) |  |
| No | 143 (36) | 11 (25) | 49 (38) | 60 (38) | 15 (35) | 8 (35) |  |
| Not reported | 73 (18) | 7 (16) | 21 (16) | 30 (19) | 8 (19) | 7 (30) |  |
| **Active smoking** |  |  |  |  |  |  | <0.001 |
| Yes, daily | 4 (1) | NA | NA | 3 (2) | 0 (0) | 1 (4) |  |
| Yes, rarely | 5 (1) | NA | NA | 3 (2) | 1 (2) | 1 (4) |  |
| Ex-smoker | 15 (4) | NA | NA | 2 (1) | 8 (19) | 5 (22) |  |
| Never smoker | 188 (47) | NA | NA | 141 (90) | 32 (74) | 15 (65) |  |
| Not reported | 185 (47) | NA | NA | 8 (5) | 2 (5) | 1 (5) |  |
| **Smoking in household** |  |  |  |  |  |  | 0.002 |
| Yes | 70 (18) | 6 (14) | 24 (18) | 34 (22) | 4 (9) | 2 (9) |  |
| No | 262 (66) | 34 (77) | 97 (75) | 89 (56) | 27 (63) | 15 (65) |  |
| Not reported | 65 (16) | 4 (9) | 9 (7) | 34 (22) | 12 (28) | 6 (26) |  |

EPIC-PCD: Ear-nose throat prospective international cohort of patients with primary ciliary dyskinesia. y: years. All characteristics are presented as N and column % with the exception of age as median and IQR: interquartile range.
NA: not applicable; age-specific questionnaire version does not include question on active smoking in this age-category. <sup>a</sup>: Chi-square test of independence.

**Figure 1:**
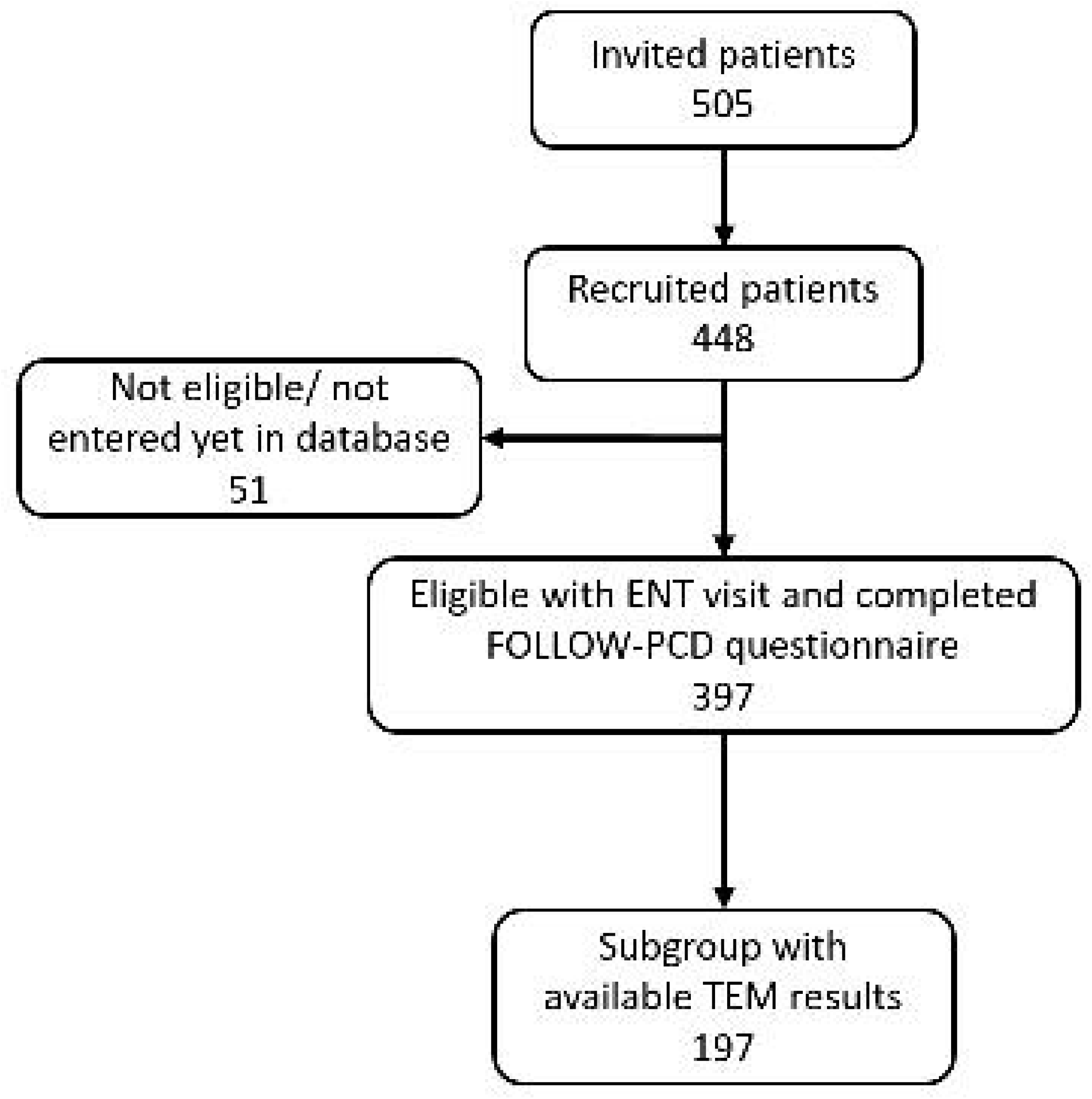
Flowchart of study population of EPIC-PCD (ENT prospective international cohort of patients with primary ciliary dyskinesia).

### Patient-reported symptoms and quality of life

In total, chronic nasal symptoms were very common; most (352; 89%) participants reported nasal symptoms during the past three months (table 2). Over half of participants (204; 52%) reported chronic nasal symptoms daily or often, which most commonly “persisted all the time” for over one-third (145; 37%) of participants. Rhinorrhoea was the most commonly (167; 47%) reported nasal symptom, although nasal discharge colour varied. Some participants reported anosmia or hyposmia (45; 11%) and nearly half (190; 48%) reported snoring. Of participants reporting snoring, 46 participants snored almost every night (12%), 83 even during periods when they did not have colds (44%). Most participants reported headaches (243; 62%), which for some occurred mainly while bending down (43; 18%). Much fewer participants (27; 7%) suffered from migraines. More ENT symptoms were reported during December when compared with other months. In comparison with children, more adults reported anosmia or hyposmia (26% vs 5%, *p* <0.005), headaches (72% vs 55%, *p* <0.110), and migraines (15% vs 2%, *p* <0.001). We did not find other differences with patient-reported sinonasal symptoms by age or sex. Only 26 (7%) participants reported no sinonasal symptoms (figure S1).

**Table 2:** Upper respiratory symptoms of past 3 months reported by EPIC-PCD participants, overall and by age group (N=397)

|  | Total<br>N (%) | Age 0-6 y<br>N (%) | Age 7-14 y<br>N (%) | Age 15-30 y<br>N (%) | Age 31-50 y<br>N (%) | Age >50 y<br>N (%) | p-value<sup>a</sup> |
| --- | --- | --- | --- | --- | --- | --- | --- |
| **Number of participants** | 397 (100) | 44 (100) | 130 (100) | 157 (100) | 43 (100) | 23 (100) |  |
| **Nasal symptoms** |  |  |  |  |  |  | 0.189 |
| Daily/often | 204 (52) | 19 (43) | 68 (52) | 72 (46) | 29 (67) | 16 (70) |  |
| Sometimes/rarely | 148 (37) | 18 (41) | 48 (37) | 65 (41) | 11 (26) | 6 (26) |  |
| Never | 45 (11) | 7 (16) | 14 (11) | 20 (13) | 3 (7) | 1 (4) |  |
| **Nasal symptoms persisting all the time<sup>b</sup>** | 145 (37) | 17 (39) | 46 (35) | 52 (33) | 22 (51) | 8 (35) | 0.786 |
| **Type of nasal symptoms<sup>c</sup>** |  |  |  |  |  |  |  |
| Rhinorrhoea | 317 (90) | 34 (92) | 102 (88) | 124 (91) | 36 (90) | 21 (95) | 0.991 |
| Blocked nose | 240 (68) | 17 (46) | 83 (72) | 96 (70) | 32 (80) | 12 (55) | 0.459 |
| Sneezing | 76 (16) | 5 (14) | 19 (16) | 32 (18) | 9 (10) | 11 (18) | 0.054 |
| Anosmia/hyposmia | 45 (13) | 0 (0) | 7 (6) | 14 (10) | 14 (35) | 10 (45) | <0.001 |
| **Colour of nasal discharge in case of rhinorrhoea<sup>d</sup>** |  |  |  |  |  |  |  |
| Clear | 65 (16) | 9 (20) | 19 (15) | 27 (17) | 6 (14) | 4 (17) | 0.940 |
| White | 60 (15) | 6 (14) | 23 (18) | 21 (14) | 6 (14) | 4 (17) | 0.922 |
| Yellow | 106 (27) | 10 (23) | 34 (26) | 45 (29) | 12 (28) | 5 (22) | 0.964 |
| Green | 75 (19) | 9 (21) | 25 (19) | 26 (27) | 9 (21) | 6 (26) | 0.907 |
| Mixed with blood | 11 (3) | 0 (0) | 1 (1) | 5 (3) | 3 (7) | 2 (9) | 0.077 |
| **Snoring** |  |  |  |  |  |  | 0.005 |
| Daily/ often | 46 (12) | 5 (11) | 14 (11) | 11 (7) | 11 (26) | 5 (22) |  |
| Sometimes/rarely | 144 (36) | 20 (46) | 51 (39) | 48 (31) | 15 (35) | 10 (43) |  |
| Never/ not reported | 207 (52) | 19 (43) | 65 (50) | 98 (62) | 17 (39) | 8 (35) |  |
| **Periods of snoring<sup>e</sup>** |  |  |  |  |  |  | 0.096 |
| Almost every night | 43 (23) | 5 (20) | 13 (20) | 9 (15) | 10 (38) | 6 (40) |  |
| Only during colds | 48 (25) | 9 (36) | 19 (29) | 11 (19) | 7 (27) | 2 (13) |  |
| Sometimes also without colds | 83 (44) | 8 (32) | 29 (45) | 35 (59) | 6 (23) | 5 (34) |  |
| Not reported | 16 (8) | 3 (12) | 4 (6) | 4 (7) | 3 (12) | 2 (13) |  |
| **Headache** |  |  |  |  |  |  | <0.001 |
| Daily/often | 50 (13) | 0 (0) | 15 (12) | 20 (13) | 9 (21) | 6 (26) |  |
| Sometimes/rarely | 193 (49) | 9 (20) | 55 (42) | 91 (58) | 28 (65) | 10 (43) |  |
| Never/ not reported | 154 (39) | 35 (80) | 60 (46) | 46 (29) | 6 (14) | 7 (30) |  |
| **Headache when bending down** | 43 (11) | 1 (2) | 7 (5) | 28 (18) | 5 (12) | 2 (9) | 0.014 |
| **Migraines** |  |  |  |  |  |  | <0.001 |
| Yes | 27 (7) | 0 (0) | 3 (2) | 10 (6) | 7 (16) | 7 (30) |  |
| No | 370 (93) | 44 (100) | 127 (98) | 147 (94) | 36 (84) | 16 (70) |  |
| **SNOT-22 completed** | 140 (35) | 14 (32) | 29 (22) | 56 (36) | 27 (63) | 14 (61) | <0.001 |
| Score median (IQR) | 31 (22-45) | 25 (15-36) | 27 (17-41) | 29 (18-40) | 36 (26-51) | 63 (35-79) | <0.001<sup>f</sup> |

EPIC-PCD: Ear-nose throat prospective international cohort of patients with primary ciliary dyskinesia. y: years. Symptoms are presented as N and column %. SNOT-22 presented as median and IQR: interquartile range. Sino Nasal Outcome Test-22 (SNOT-22) questionnaire on chronic rhinosinusitis related items scored 0-5 ("No problem" to "Problem as bad as it can be"), total score range 0-110, mild 0-20, moderate 21-50, severe ≥51. <sup>a</sup>: Chi-square test of independence. <sup>b</sup>: Among 352 people with chronic
nasal symptoms <sup>c</sup>: Among 352 people with chronic nasal symptoms, categories are not exclusive. <sup>d</sup>: Among 317 people with rhinorrhea, categories are not exclusive. <sup>e</sup>: Among 190 people with snoring. <sup>f</sup>: Kruskal-Wallis test.

In total, 140 (35%) participants who were most commonly adults completed SNOT-22 questionnaires. The median score was 31 (IQR 22–45), reflecting a moderate effect of CRS symptoms on QoL (table 2). Median SNOT-22 scores were higher with age; we observed the most severe effect on QoL (63; IQR 35–79) among participants aged 50 years and older (*p* <0.001), and among participants with daily nasal symptoms (figure S2).

### Sinonasal clinical examinations

We excluded 15 of 397 study participants from our analysis of sinonasal examination findings for remote sinonasal consultations without sinonasal examinations. Among the remaining 382 participants, recording of sinonasal findings from sinonasal examinations was incomplete for some (table 3). For 165 (43%) participants, the nose appeared blocked, while nasal discharge was mainly serous (87; 30%) or sero-mucous (122; 43%). Abnormal nasal mucosa findings were recorded for 169 (44%), specifically mucosal oedema for 108 (26%) participants. Nasal polyps were assessed in 353 participants, and identified among 52 (14%; median age 20 years; IQR 14–36) participants with 19 (37%) located bilaterally. Of the 52 participants, 39 (75%) had nasal polyps either partially (9; 17%) or fully (9; 17%) blocking nasal passages. Nasal turbinates were hypertrophic in 129 participants (34%) and 118 participants had deviated septum (31%). 51 participants had facial pain (13%) at examination. When compared with children, more adults had nasal polyps, hypertrophic turbinates, deviated septum, and facial pain (all *p* <0.003). We did not find differences according to sex.

**Table 3:** Sinonasal examination results of EPIC-PCD participants, overall and by age group (N=382)

|  | Total N (%) | Age 0-6 y<br>N (%) | Age 7-14 y<br>N (%) | Age 15-30 y<br>N (%) | Age 31-50 y<br>N (%) | Age >50 y<br>N (%) | p-value<sup>a</sup> |
| --- | --- | --- | --- | --- | --- | --- | --- |
| **ENT consultations on site** | 382 (100) | 38 (100) | 123 (100) | 155 (100) | 43 (100) | 23 (100) |  |
| **Nose appearance** |  |  |  |  |  |  | 0.054 |
| Normal | 209 (55) | 23 (60) | 76 (62) | 85 (55) | 16 (37) | 9 (39) |  |
| Blocked | 165 (43) | 14 (37) | 45 (37) | 68 (44) | 26 (61) | 12 (52) |  |
| Not recorded | 8 (2) | 1 (3) | 2 (2) | 2 (1) | 1 (2) | 2 (9) |  |
| **Nasal discharge present** |  |  |  |  |  |  | 0.767 |
| Yes | 286 (75) | 28 (74) | 94 (76) | 112 (72) | 34 (79) | 18 (78) |  |
| No | 88 (23) | 9 (24) | 27 (22) | 41 (27) | 7 (16) | 4 (18) |  |
| Not recorded | 8 (2) | 1 (2) | 2 (2) | 2 (1) | 2 (5) | 1 (4) |  |
| **Type of nasal discharge<sup>b</sup>** |  |  |  |  |  |  | 0.866 |
| Serous | 87 (30) | 10 (36) | 30 (32) | 32 (28) | 9 (27) | 6 (33) |  |
| Sero-mucous | 122 (43) | 11 (39) | 42 (45) | 46 (41) | 16 (47) | 7 (39) |  |
| Muco-purulent | 66 (23) | 6 (21) | 19 (20) | 29 (26) | 8 (23) | 4 (22) |  |
| Mixed with blood | 3 (1) | 1 (4) | 0 (0) | 1 (1) | 0 (0) | 1 (6) |  |
| Not recorded | 8 (3) | 0 (0) | 3 (3) | 4 (4) | 1 (3) | 0 (0) |  |
| **Nasal mucosa** |  |  |  |  |  |  | 0.018 |
| Abnormal | 169 (44) | 13 (34) | 53 (43) | 68 (44) | 21 (49) | 14 (61) |  |
| Normal | 202 (53) | 21 (55) | 67 (55) | 86 (55) | 21 (49) | 7 (30) |  |
| Not recorded | 11 (3) | 4 (11) | 3 (2) | 1 (1) | 1 (2) | 2 (9) |  |
| **Nasal polyps** |  |  |  |  |  |  | 0.001 |
| Yes | 52 (14) | 2 (5) | 11 (9) | 22 (14) | 12 (28) | 5 (22) |  |
| No | 301 (79) | 28 (74) | 100 (81) | 127 (82) | 29 (68) | 17 (74) |  |
| Not assessed | 29 (7) | 8 (21) | 12 (10) | 6 (4) | 2 (5) | 1 (4) |  |
| **Nasal polyps size<sup>cd</sup>** |  |  |  |  |  |  | 0.827 |
| Fully blocking | 9 (17) | 1 (50) | 3 (27) | 6 (4) | 2 (17) | 0 (0) |  |
| Partially blocking | 39 (75) | 1 (50) | 7 (64) | 17 (77) | 9 (75) | 5 (100) |  |
| Not assessed | 4 (8) | 0 (0) | 1 (9) | 2 (9) | 1 (8) | 0 (0) |  |
| **Bilaterally<sup>cd</sup>** |  |  |  |  |  |  | 0.263 |
| Fully blocking | 17 (33) | 1 (50) | 3 (27) | 6 (4) | 4 (33) | 3 (60) |  |
| Partially blocking | 2 (4) | 0 (0) | 2 (18) | 0 (0) | 0 (0) | 0 (0) |  |
| Not recorded | 33 (64) | 1 (50) | 6 (55) | 16 (72) | 8 (67) | 2 (40) |  |
| **Unilaterally<sup>cd</sup>** |  |  |  |  |  |  | 0.213 |
| Fully blocking | 39 (75) | 2 (5) | 7 (6) | 15 (10) | 11 (26) | 4 (17) |  |
| Partially blocking | 4 (8) | 0 (0) | 3 (2) | 1 (1) | 0 (0) | 0 (0) |  |
| Not recorded | 9 (17) | 0 (0) | 1 (9) | 6 (27) | 1 (8) | 1 (20) |  |

EPIC-PCD: Ear-nose throat prospective international cohort of patients with primary ciliary dyskinesia. y: years. Symptoms are presented as N and column %. <sup>a</sup>: Chi-square test of independence. <sup>b</sup>: Among 286 people with nasal discharge. <sup>c</sup>: Among 52 participants with nasal polyps. <sup>d</sup>: Nasal polyps described as partially blocking or with Lildholdt score 1 or 2, fully blocking or with Lildholdt score 3.

**Table 3 (continued):** Sinonasal examination results of EPIC-PCD participants, overall and by age group (N=382)
|  | Total N (%) | Age 0-6 y<br>N (%) | Age 7-14 y<br>N (%) | Age 15-30 y<br>N (%) | Age 31-50 y<br>N (%) | Age >50 y<br>N (%) | p-value<sup>a</sup> |
| --- | --- | --- | --- | --- | --- | --- | --- |
| **ENT consultations on site** | 382 (100) | 38 (100) | 123 (100) | 155 (100) | 43 (100) | 23 (100) |  |
| **Inferior nasal turbinates** |  |  |  |  |  |  | 0.003 |
| Normal | 221 (58) | 21 (55) | 67 (55) | 100 (64) | 21 (49) | 12 (52) |  |
| Hypertrophy | 129 (34) | 13 (34) | 47 (38) | 46 (30) | 15 (35) | 8 (35) |  |
| Atrophy | 3 (1) | 0 (0) | 0 (0) | 0 (0) | 3 (7) | 0 (0) |  |
| Not recorded | 29 (7) | 4 (11) | 9 (7) | 9 (6) | 4 (9) | 3 (13) |  |
| **Deviated nasal septum** |  |  |  |  |  |  | <0.001 |
| Yes | 118 (31) | 2 (5) | 28 (23) | 66 (42) | 12 (28) | 10 (43) |  |
| Bulging forward | 5 (1) | 1 (3) | 0 (0) | 1 (1) | 2 (5) | 1 (5) |  |
| No | 235 (62) | 28 (74) | 88 (71) | 81 (52) | 28 (65) | 10 (43) |  |
| Not recorded | 24 (6) | 7 (18) | 7 (6) | 7 (5) | 1 (2) | 2 (9) |  |
| **Facial pain or sensitivity** |  |  |  |  |  |  | <0.001 |
| Yes | 51 (13) | 0 (0) | 9 (7) | 21 (14) | 12 (28) | 9 (39) |  |
| No | 302 (79) | 27 (71) | 107 (87) | 127 (82) | 28 (65) | 13 (57) |  |
| Not recorded | 29 (8) | 11 (29) | 7 (6) | 7 (4) | 3 (7) | 1 (4) |  |

EPIC-PCD: Ear-nose throat prospective international cohort of patients with primary ciliary dyskinesia. y: years. Symptoms are presented as N and column %. <sup>a</sup>: Chi-square test of independence

### Information on management of upper airways

At baseline, 76 (19%) participants had hospitalisations since previous consultation, yet it was unattributed to upper respiratory infections (table S2). A small proportion of participants (17; 4%) underwent elective operations, 9 of them for sinonasal complications (53%), during this period.

Nearly one-quarter (83; 24%) of 339 participants were prescribed nasal corticosteroids, most commonly for year-round use (70; 84%). Out of 203 participants, the most common relevant nasal corticosteroid instruction involved regular nose blowing (170; 84%); out of 208 participants, instructions commonly involved nasal rinsing (190; 91%)—both instructions recommended mostly year-round use. Lastly, 50 (19%) of 269 participants (19%) were prescribed upper airway nebulisation prescriptions with isotonic saline (17; 34%), hypertonic saline (23; 46%), or other medication (9; 18%). Most commonly (47; 95%) prescribed using year-round.

### Factors associated with sinonasal disease

We found age 10 years and older associated with higher risk of sinonasal disease, more profoundly when comparing participants ages 31–40 years with ages 0–10 years (odds ratio OR: 14.29, 95% CI: 5.17–39.48). Even after accounting for age, risk also differed based on study centre (table S3). We did not find associations with sex, tobacco smoke exposure, or season when consultations took place (figure 2). In the subgroup analyses of 197 participants with available TEM results (table S1), we found no association between ciliary ultrastructural defect class and risk of sinonasal disease (figure S3).

**Figure 2:**
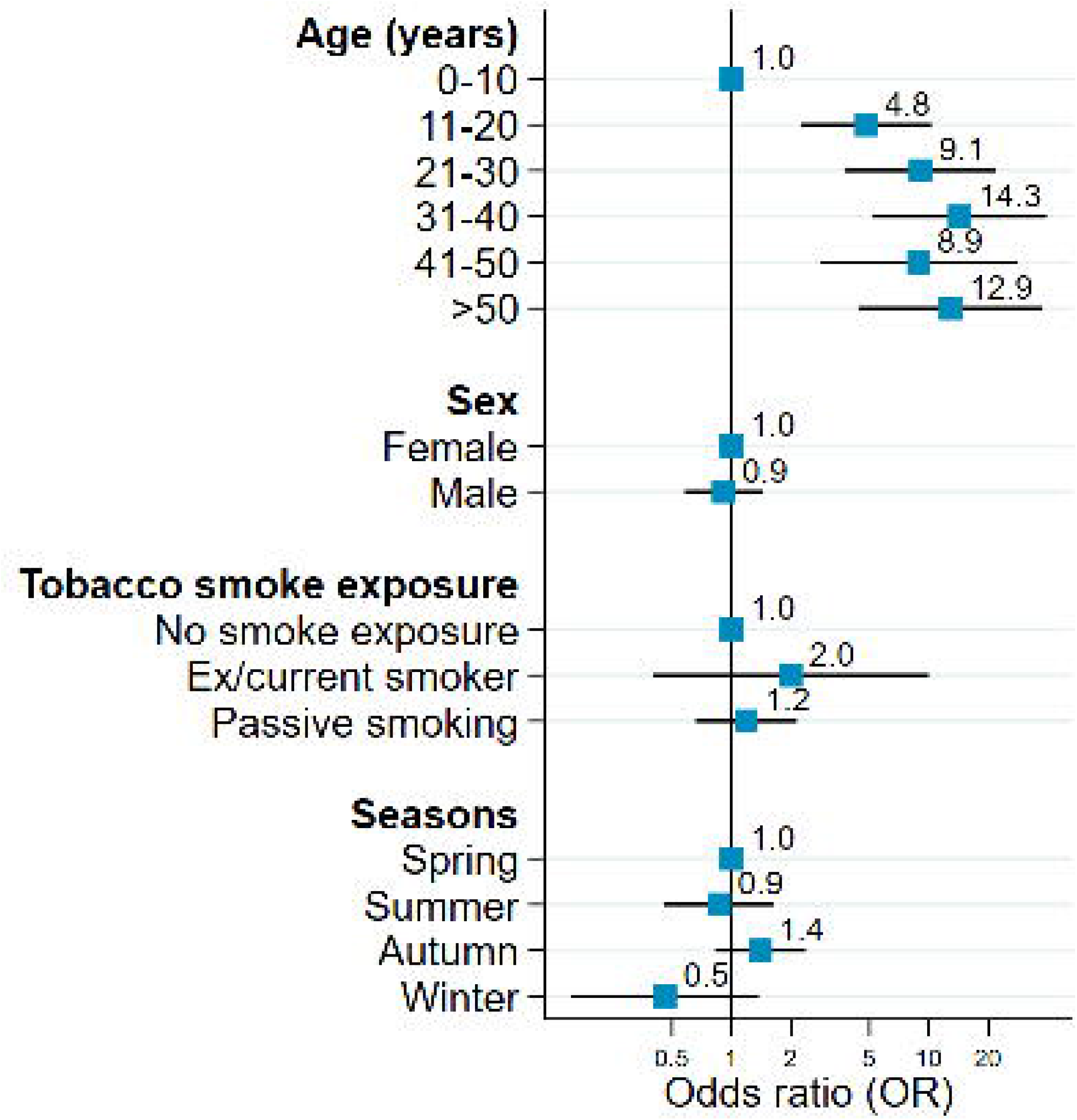
Factors associated with sinonasal disease in EPIC-PCD participants (N=397). Sinonasal disease defined by composite outcome score consisting of three variables: patient-reported headache while bending down as a proxy for sinusitis, and ENT examination findings of nasal polyps and facial pain. EPIC-PCD: Ear-nose throat prospective international cohort of patients with primary ciliary dyskinesia. Odds ratio (OR) indicated by squares and 95% confidence intervals by horizontal lines.

## Discussion

Our study benefitted from data from the first prospective, multicentre, international ENT cohort of patients with PCD. Even though we performed our study during the COVID-19 pandemic, with much lower prevalence of viral infections, most participants reported chronic nasal symptoms, most commonly rhinorrhea. Our results showed sinonasal symptoms and clinical examination findings indicated chronic inflammation that is also more common with increasing age. Overall, QoL— measured by SNOT-22—was moderately affected by CRS (median score 31; IQR 22–45).

Anterior rhinoscopic or endoscopic findings, such as nasal polyps, hypertrophic turbinates, and deviated septum, as well as facial pain at examination, were more commonly found among adults than children. We found the risk of sinonasal disease increased with age and associated with study centre.

### Strengths and limitations

Our study’s main strength includes our use of data from a large, prospective, international cohort with an overall recruitment rate of 89%. We are the first to describe patient-reported sinonasal symptoms and sinonasal examination findings obtained during the same consultation for PCD. Another strength is our use of FOLLOW-PCD—it allowed standardised records of disease-specific information and comparisons between participating centres. We excluded participants if their data were not yet entered in the study database or they did not meet eligibility criteria. We have no reasons to think exclusions were not random or affected the representation of our study population, yet participants with more sinonasal symptoms might be more willing to join EPIC-PCD when invited. We expect small risks of recall bias for patient-reported symptoms since questionnaires ask about the last three months. However, these symptoms are unspecific and part of participants’ daily life, so they might be underreported. Particularly, assessing anosmia or hyposmia among young children is difficult and possibly underreported by parents. Mostly, adult participants completed SNOT-22 questionnaires, which is expected since it is only validated for adults and not used at all participating centres. We still do not know if the score sufficiently capture or underestimate effects from CRS on QoL among children. Another reason might be that the prevalence of CRS increases with age among adults in the general population; in particular CRS without nasal polyps is more prevalent among adults 40 years or younger and CRS with nasal polyps is more evident among adults 40 years or older [24]. Although our cohort was set up at the beginning of the COVID-19 pandemic, based on data from the COVID-PCD study SARS-CoV-2 infections were infrequent and caused generally mild to moderate symptoms among people with PCD [32], probably due to participants’ careful shielding behaviours [33]. Generally, people with PCD and their shielding behaviours likely led to fewer infections, resulted in lower prevalence, and underestimated sinonasal problems, yet almost all participants reported nasal symptoms.

### Comparison with other studies

Previous studies of upper respiratory symptoms also showed ENT symptoms common among people with PCD; however, ENT symptom definitions varied, making comparisons difficult. For instance, in a prospective study using a nationwide survey based on the FOLLOW-PCD questionnaire in Switzerland, 70 (94%) of 74 participants reported chronic nasal symptoms with rhinorrhoea (65%), blocked nose (55%), or anosmia (38%) [9]. In comparison with this study, an older population or differences in upper airway management, could explain higher prevalence of symptoms. Differences in upper airway management among participating study centres might explain the risk of sinonasal disease differences we found, yet in depth comparisons require more detailed data. Similar to our findings, a prospective study in North America described CRS among 47 children with nasal polyps (3; 6%), snoring (23; 49%), and a mean SNOT-22 score of 36.4 [34]. In a retrospective study in France, 63 of 64 adults reported sinonasal problems along with pathological nasal endoscopic findings [35], which is similar to our adult population’s chronic nasal problems. In the same study, there was no correlation of ENT disease severity with ciliary ultrastructural defects. In another study assessing 39 adults with PCD and CRS in Italy, 59% had nasal polyps and more severely affected QoL—measured by SNOT-22 score—than those without nasal polyps [36]. Their findings were more severe than among our population, probably because we included children and more young adults. A study including 67 adults with PCD in Japan supports our finding that nasal polyps were observed more frequently with increasing age [37]. Although we did not observe similarly, higher odds of having CRS have been described for tobacco smoke exposure in the general population. However, our population reported a small number of participants exposed to tobacco, particularly active smoking [38].

### Conclusion

We found sinonasal problems persist throughout life among people with PCD. In particular, more adults had nasal polyps and reported anosmia or hyposmia, showing complications of CRS increase with age, possibly due to ongoing chronic inflammation. Although most participants frequently reported sinonasal symptoms, not all were prescribed sinonasal treatment or management, which could be due to patients underreporting or lack of standardised care and evidence-based PCD management guidelines for upper airways. Our study reinforces the importance of regular sinonasal examinations for PCD patients of all ages and the need to develop evidence-based sinonasal treatments as part of the overall PCD management.

## Data Availability

The datasets used and analysed during the current study are available from the study PI Dr Myrofora Goutaki upon reasonable request.

## Author contribution

M Goutaki developed the concept and designed the study. M Goutaki, YT Lam manage the study. YT Lam cleaned and standardised the data and performed statistical analyses supervised by M Goutaki. YT Lam and M Goutaki drafted the manuscript. All authors commented and revised the manuscript. YT Lam and M Goutaki take final responsibility for the content.

## Statement on funding sources and conflicts of interest

This study is funded by a Swiss National Science Foundation Ambizione fellowship (PZ00P3_185923). The authors participate in the BEAT-PCD (Better experimental approaches to treat PCD) clinical research collaboration, supported by the European Respiratory Society, and most centres are members of the PCD core of ERN-LUNG (European Reference Network on rare respiratory diseases). P Latzin received grants or honoraria for participation in data safety monitoring boards or advisory boards from Vertex, Vifor, OM Pharma, Polyphor, Santhera (DMC), and Sanofi Aventis within the last 36 months. J Roehmel received grants, clinical study recompensations from Vertex, INSMED, Medical Research Council/UK, BMBF, Mukoviszidose Institut.

## Acknowledgements

We thank all people with PCD and their families participating in EPIC-PCD and PCD support organisations (especially PCD Family Support Group UK; Association ADCP France; Kartagener Syndrom und Primäre Ciliäre Dyskinesie e. V. Deutschland/ Deutschschweiz; Asociación Nacional de Pacientes con Discinesia Ciliar Primaria DCP España/PCD Spain) for their close collaboration. We also thank all researchers from the participating centres involved in enrolment, data collection, and data entry who work closely with us (listed below as collaborators of the EPIC-PCD study). We are grateful for everyone who contributed to translations of the FOLLOW-PCD questionnaire in Dutch, Flemish, French, Norwegian, Spanish, and Turkish. We thank Kristin Marie Bivens (ISPM, University of Bern) for her editorial assistance.

## Collaborators of the EPIC-PCD study

(listed in alphabetical order): Dilber Ademhan (Hacettepe University, Turkey), Mihaela Alexandru (AP-HP, France), Andreas Anagiotos (Nicosia General Hospital, Cyprus), Miguel Armengot (La Fe University and Polytechnic Hospital, Spain), Lionel Benchimol (University Hospital of Liège, Belgium), Achim G Beule (University of Münster, Germany), Irma Bon (Vrije Universiteit, the Netherlands), Mieke Boon (University Hospital Leuven, Belgium), Marina Bullo (University of Bern, Switzerland), Andrea Burgess (University of Southampton, UK), Doriane Calmes (University Hospital of Liège, Belgium), Carmen Casaulta (University of Bern, Switzerland), Marco Caversaccio (University of Bern, Switzerland), Nathalie Caversaccio (University of Bern, Switzerland), Bruno Crestani (RESPIRARE, France), Suzanne Crowley (University of Oslo, Norway), Sinan Ahmed. D. Dheyauldeen (University of Oslo, Norway), Sandra Diepenhorst (Vrije Universiteit, The Netherlands), Nagehan Emiralioglu (Hacettepe University, Turkey), Ela Erdem Eralp (Marmara University, Turkey), Pinar Ergenekon (Marmara University, Turkey), Nathalie Feyaerts (University Hospital Leuven, Belgium), Gavriel Georgiou (Nicosia General Hospital, Cyprus), Amy Glen (University of Southampton, UK), Christine van Gogh (Vrije Universiteit Amsterdam, the Netherlands), Yasemin Gokdemir (Marmara University, Turkey), Myrofora Goutaki (University of Bern, Switzerland), Onder Gunaydın (Hacettepe University, Turkey), Eric G Haarman (Vrije Universiteit Amsterdam, The Netherlands), Amanda Harris (University of Southampton, UK), Isolde Hayn (Charité-Universitätsmedizin Berlin, Germany), Simone Helms (University of Münster, Germany), Sara-Lynn Hool (University of Bern, Switzerland), Isabelle Honoré (RESPIRARE, France), Hasnaa Ismail Koch (University of Southampton, UK), Bülent Karadag (Marmara University, Turkey), Céline Kempeneers (University Hospital of Liège, Belgium), Synne Kennelly (University of Oslo, Norway), Elisabeth Kieninger (University of Bern, Switzerland), Sookyung Kim (AP-HP, France), Panayiotis Kouis (University of Cyprus, Cyprus), Yin Ting Lam (University of Bern, Switzerland), Philipp Latzin (University of Bern, Switzerland), Marie Legendre (RESPIRARE, France), Natalie Lorent (University Hospital Leuven, Belgium), Jane S Lucas (University of Southampton, UK), Bernard Maitre (RESPIRARE, France), Alison McEvoy (University of Southampton, UK), Rana Mitri-Frangieh (RESPIRARE, France), David Montani (RESPIRARE, France), Loretta Müller (University of Bern, Switzerland), Noelia Muñoz (La Fe University and Polytechnic Hospital, Spain), Heymut Omran (University of Münster, Germany), Ugur Ozcelik (Hacettepe University, Turkey), Beste Ozsezen (Hacettepe University, Turkey), Samantha Packham (University of Southampton, UK), Jean-François Papon (AP-HP, France), Clara Pauly (University Hospital of Liège, Belgium), Charlotte Pioch (Charité-Universitätsmedizin Berlin, Germany), Anne-Lise ML Poirrier (University Hospital of Liège, Belgium), Johanna Raidt (University of Münster, Germany), Ana Reula (La Fe University, Spain), Rico Rinkel (Vrije Universiteit Amsterdam, the Netherlands), Jobst Roehmel (Charité-Universitätsmedizin Berlin, Germany), Andre Schramm (University of Münster, Germany), Simone Tanner (Vrije Universiteit, the Netherlands), Guillaume Thouvenin (RESPIRARE, France), Woolf T Walker (University of Southampton, UK), Hannah Wilkins (University of Southampton, UK), Panayiotis Yiallouros (University of Cyprus, Cyprus), Ali Cemal Yumusakhuylu (Marmara University, Turkey), Niklas Ziegahn (Charité-Universitätsmedizin Berlin, Germany)

## Figure legends

**Figure S1:** Proportion of patient-reported symptoms (snoring, headache, nasal symptoms) during the past three months among EPIC-PCD participants (N=397).

EPIC-PCD: Ear-nose throat prospective international cohort of patients with primary ciliary dyskinesia.

**Figure S2:** Sino-Nasal Outcome Test (SNOT)-22 score by frequency of patient-reported nasal symptoms during the past three months among EPIC-PCD participants (N=140).

SNOT-22 scores 0–5 (“No problem” to “Problem as bad as it can be”); total score range 0–110; mild 0–20, moderate 21–50; severe ≥ 51. Score ranges indicated by horizontal lines. EPIC-PCD: Ear-nose throat prospective international cohort of patients with primary ciliary dyskinesia.

**Figure S3:** Association of ciliary ultrastructural defect with sinonasal disease in EPIC-PCD participants (N=197).

Sinonasal disease defined by composite outcome score consisting of three variables: patient-reported headache while bending down as a proxy for sinusitis, ENT examination findings of nasal polyps, and facial pain. EPIC-PCD: Ear-nose throat prospective international cohort of patients with primary ciliary dyskinesia. Odds ratio (OR) indicated by squares and 95% confidence intervals (CI) by horizontal lines.

## Notes

### Clinical Trial

NCT04611516

### Clinical Protocols

https://bmjopen.bmj.com/content/11/10/e051433.long

