## Supplement material for "Sinonasal disease among patients with primary ciliary dyskinesia – an international study"

**Table S1**: Results of transmission electron microscopy of EPIC-PCD participants, overall and by age group (N=197)

| | **Total N (%)** | **Age 0-6 y N (%)** | **Age 7-14 y N (%)** | **Age 15-30 y N (%)** | **Age 31-50 y N (%)** | **Age >50 y N (%)** |
|---|---|---|---|---|---|---|
| **Number of participants** | 197 (100) | 22 (100) | 66 (100) | 69 (100) | 25 (100) | 15 (100) |
| ODA & IDA-defect | 62 (31) | 10 (45) | 17 (26) | 27 (40) | 6 (24) | 2 (13) |
| ODA-defect | 27 (13) | 1 (5) | 12 (18) | 10 (14) | 2 (8) | 2 (13) |
| Microtubular disorganisation & IDA defect | 29 (15) | 2 (9) | 10 (15) | 10 (14) | 2 (8) | 5 (34) |
| Central complex defect | 17 (9) | 0 (0) | 3 (4) | 6 (9) | 7 (28) | 1 (7) |
| Normal ultrastructure | 41 (21) | 9 (41) | 15 (23) | 9 (13) | 5 (20) | 3 (20) |
| Non-hallmark defect | 21 (11) | 0 (0) | 9 (14) | 7 (10) | 3 (12) | 2 (13) |

EPIC-PCD: Ear-nose throat prospective international cohort of patients with primary ciliary dyskinesia. y: years. Results presented as N and column %.

**Table S2**: Upper airway management of EPIC-PCD participants, overall and by age group (N=397)

| | Total N (%) | Age 0-6 y N (%) | Age 7-14 y N (%) | Age 15-30 y N (%) | Age 31-50 y N (%) | Age >50 y N (%) | *p*-value[a] |
|---|---|---|---|---|---|---|---|
| **Number of participants** | 397 (100) | 44 (100) | 130 (100) | 157 (100) | 43 (100) | 23 (100) | |
| **Hospitalisation since last consultation** | 76 (19) | 7 (16) | 17 (13) | 25 (16) | 18 (42) | 9 (39) | <0.001 |
| Not recorded | 321 (81) | 37 (84) | 113 (87) | 132 (84) | 25 (58) | 14 (61) | |
| For sinonasal surgeries | 9 (2) | 0 (0) | 3 (2) | 2 (1) | 3 (7) | 1 (4) | 0.197 |
| For other surgeries | 11 (3) | 0 (0) | 2 (2) | 4 (3) | 0 (0) | 5 (22) | <0.001 |
| **Antibiotics prescribed for infection of** | | | | | | | 0.016 |
| upper respiratory tract | 19 (5) | 1 (2) | 8 (6) | 10 (6) | 0 (0) | 0 (0) | |
| lower respiratory tract | 71 (18) | 9 (20) | 27 (21) | 23 (15) | 9 (21) | 3 (13) | |
| other | 20 (5) | 2 (5) | 3 (2) | 5 (3) | 6 (14) | 4 (17) | |
| Not recorded | 287 (72) | 32 (73) | 92 (71) | 119 (76) | 28 (65) | 16 (70) | |
| **Nasal corticosteroids** | | | | | | | <0.001 |
| Yes | 83 (21) | 3 (7) | 19 (15) | 35 (22) | 17 (40) | 9 (39) | |
| No | 256 (64) | 33 (75) | 98 (75) | 103 (66) | 18 (41) | 4 (17) | |
| Not recorded | 58 (15) | 8 (18) | 13 (10) | 19 (12) | 8 (19) | 10 (44) | |
| During all year | 70 (18) | 1 (2) | 17 (13) | 3 (20) | 13 (30) | 8 (35) | <0.001 |
| During exacerbations | 11 (3) | 2 (5) | 2 (2) | 4 (3) | 3 (7) | 0 (0) | 0.353 |
| **Nasal rinsing** | | | | | | | <0.001 |
| Yes | 190 (48) | 11 (25) | 65 (50) | 81 (51) | 23 (54) | 10 (34) | |
| No | 118 (30) | 20 (45) | 50 (39) | 42 (27) | 4 (9) | 2 (9) | |
| Not recorded | 89 (22) | 13 (30) | 15 (11) | 34 (22) | 16 (37) | 11 (47) | |
| During all year | 156 (39) | 10 (23) | 57 (44) | 64 (41) | 17 (40) | 8 (35) | 0.536 |
| During exacerbations | 21 (5) | 1 (2) | 5 (4) | 10 (6) | 3 (7) | 2 (9) | 0.692 |
| **Regular nose blowing** | | | | | | | <0.001 |
| Yes | 170 (43) | 17 (39) | 66 (51) | 56 (36) | 21 (49) | 10 (44) | |
| No | 123 (31) | 12 (27) | 45 (35) | 60 (38) | 5 (12) | 1 (4) | |
| No recorded | 104 (26) | 15 (34) | 19 (15) | 41 (26) | 17 (40) | 12 (52) | |
| During all year | 154 (39) | 16 (36) | 64 (49) | 49 (31) | 17 (40) | 8 (35) | 0.361 |
| During exacerbations | 6 (2) | 0 (0) | 1 (1) | 4 (3) | 1 (2) | 0 (0) | 0.600 |
| **Nebulisation** | | | | | | | <0.001 |
| Yes | 50 (13) | 2 (5) | 19 (15) | 16 (10) | 10 (23) | 3 (13) | |
| No | 219 (55) | 26 (59) | 84 (64) | 91 (58) | 12 (28) | 6 (26) | |
| Not recorded | 128 (32) | 16 (36) | 27 (21) | 50 (32) | 21 (49) | 14 (61) | |
| During all year | 47 (12) | 2 (5) | 18 (14) | 16 (10) | 8 (19) | 3 (13) | 0.402 |
| During exacerbations | 0 (0) | 0 (0) | 0 (0) | 0 (0) | 0 (0) | 0 (0) | |
| Isotonic saline | 17 (4) | 0 (0) | 6 (5) | 4 (3) | 6 (14) | 1 (4) | 0.024 |
| Hypertonic saline | 23 (6) | 2 (5) | 8 (6) | 9 (6) | 3 (7) | 1 (4) | 0.989 |

EPIC-PCD: Ear-nose throat prospective international cohort of patients with primary ciliary dyskinesia. y: years. Isotonic saline: NaCl 0.9%. Hypertonic saline: NaCl >0.9%. [a]: Chi-square test of independence.

**Table S3**: Association of EPIC-PCD study centre with sinonasal disease (N=397)

| | Number of participants (%) | Odds ratio | 95% confidence interval |
|---|---|---|---|
| **Study centres** | 397 (100) | | |
| Paris | 53 (13) | **Reference** | |
| Amsterdam | 26 (7) | 0.07 | 0.02–3.33 |
| Ankara | 60 (15) | 0.22 | 0.09–0.49 |
| Berlin | 43 (11) | 0.48 | 0.21–1.08 |
| Bern | 7 (2) | 0.00 | 0.00–0.00 |
| Cyprus | 22 (5) | 1.11 | 0.43–2.87 |
| Istanbul | 66 (17) | 0.36 | 0.17–0.76 |
| Leuven | 12 (3) | 0.89 | 0.29–2.78 |
| Liège | 10 (3) | 1.35 | 0.38–4.86 |
| Oslo | 39 (10) | 0.25 | 0.10–0.63 |
| Southampton | 43 (10) | 0.11 | 0.04–0.32 |
| Valencia | 16 (4) | 0.21 | 0.05–0.81 |

Results of univariable ordinal logistic regression model, including only study centre as an explanatory variable (reference centre: Paris). Sinonasal disease defined with composite outcome score consisting of three variables: patient-reported headache while bending down as a proxy for sinusitis, and ENT examination findings of nasal polyps and facial pain. EPIC-PCD: Ear-nose throat prospective international cohort of patients with primary ciliary dyskinesia.

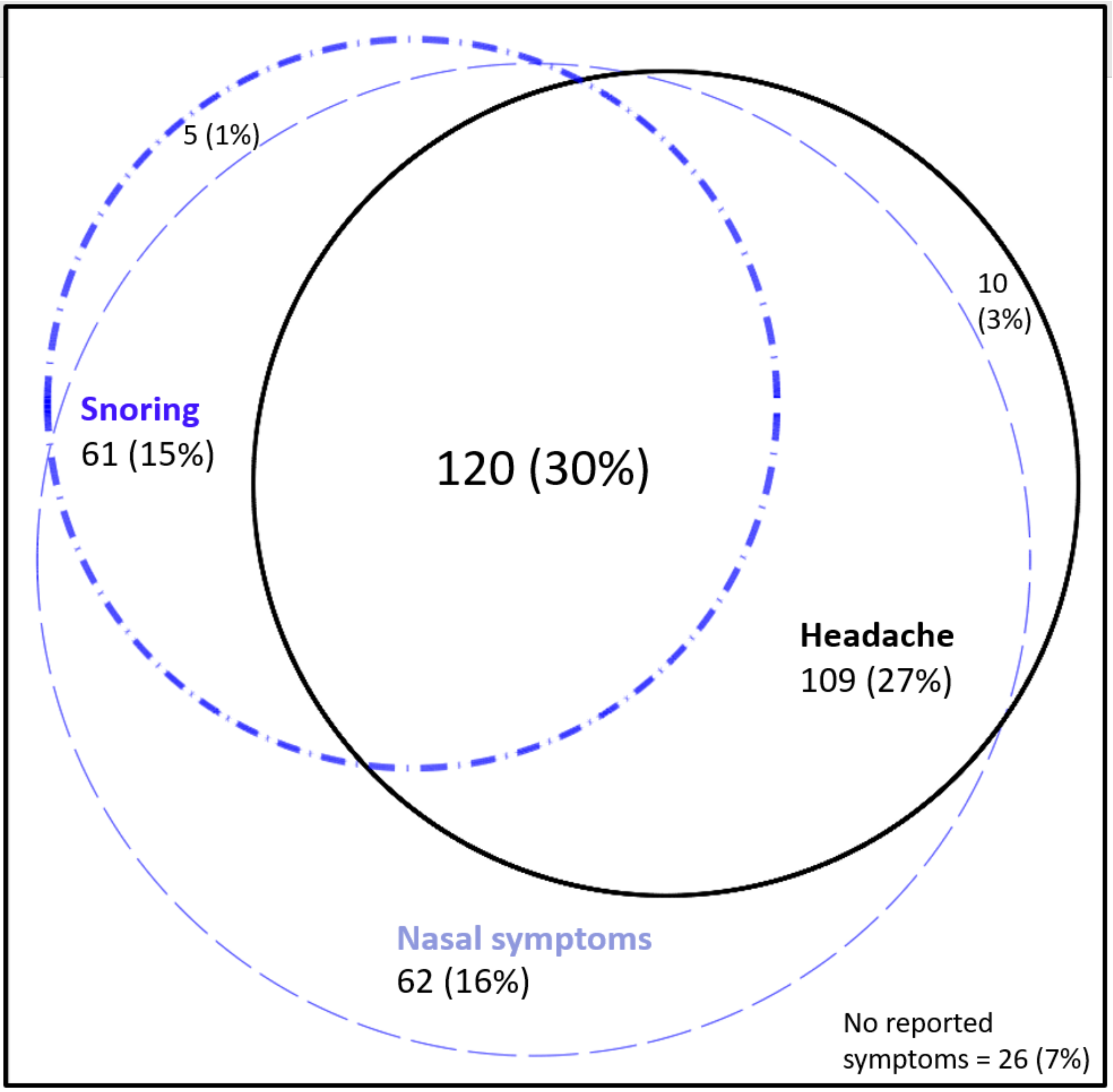

**Figure S1:** Proportion of patient-reported symptoms (snoring, headache, nasal symptoms) during the past three months among EPIC-PCD participants (N=397).

EPIC-PCD: Ear-nose throat prospective international cohort of patients with primary ciliary dyskinesia.

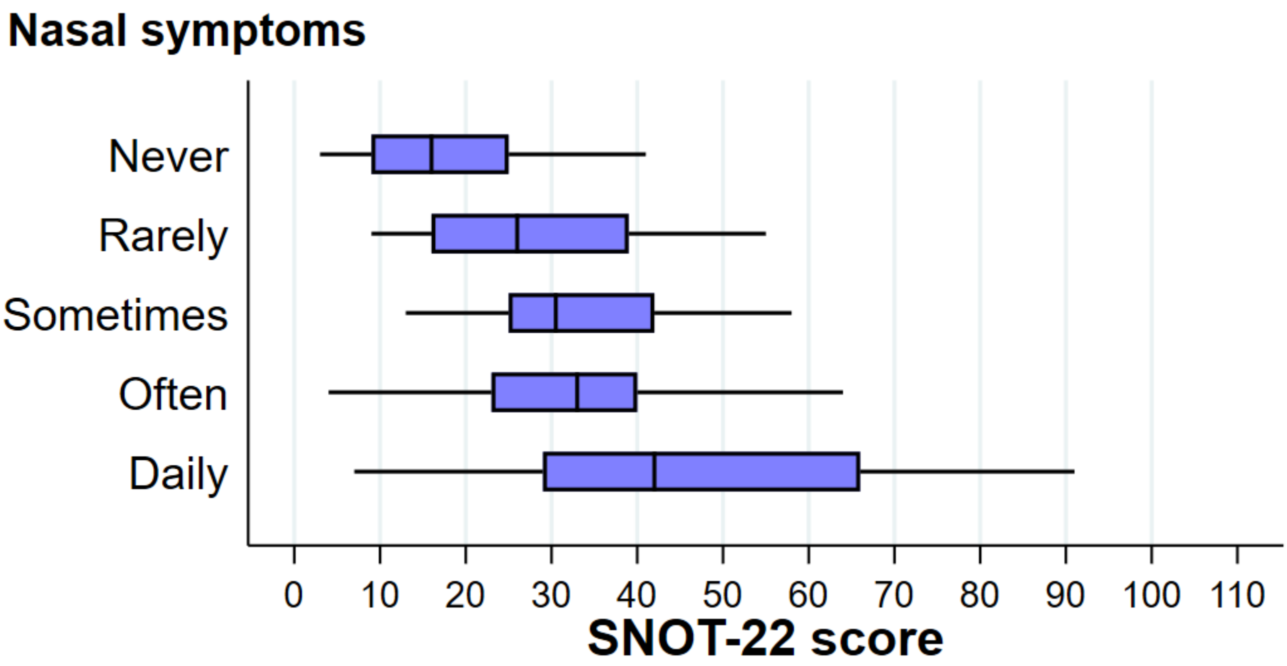

**Figure S2:** Sino-Nasal Outcome Test (SNOT)-22 score by frequency of patient-reported nasal symptoms during the past three months among EPIC-PCD participants (N=140).

SNOT-22 scores 0–5 (“No problem” to “Problem as bad as it can be”); total score range 0–110; mild 0–20, moderate 21–50; severe ≥ 51. Score ranges indicated by horizontal lines. EPIC-PCD: Ear-nose throat prospective international cohort of patients with primary ciliary dyskinesia.

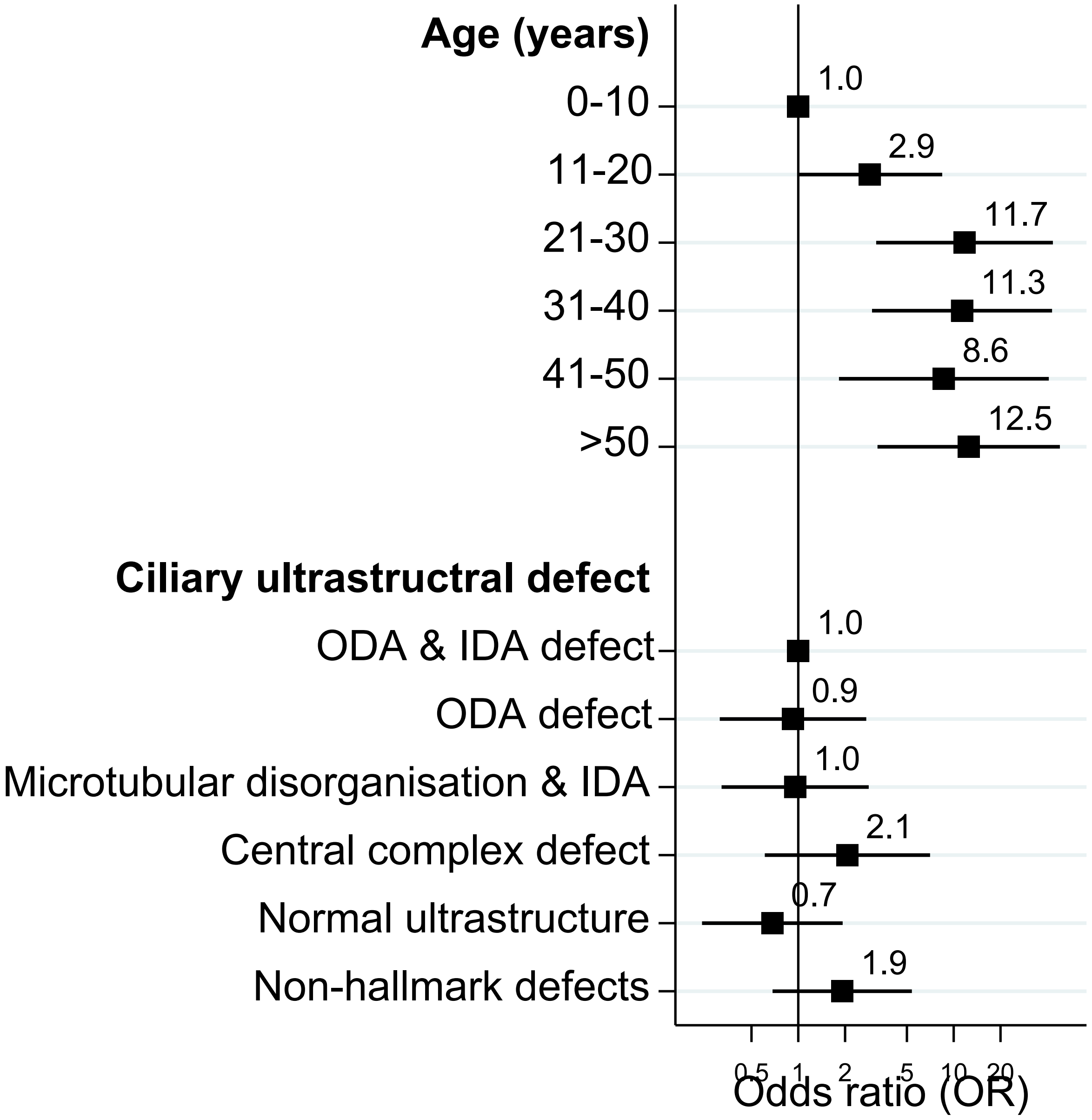

**Figure S3:** Association of ciliary ultrastructural defect with sinonasal disease in EPIC-PCD participants (N=197).

Sinonasal disease defined by composite outcome score consisting of three variables: patient-reported headache while bending down as a proxy for sinusitis, ENT examination findings of nasal polyps, and facial pain. EPIC-PCD: Ear-nose throat prospective international cohort of patients with primary ciliary dyskinesia. Odds ratio (OR) indicated by squares and 95% CI indicated by horizontal lines.
